# Burden of Cancer in Kilifi County: A 10-Year Retrospective Descriptive Study

**DOI:** 10.64898/2026.05.20.26353643

**Authors:** Moses Masha, Regina Wachuka Mbugua, Mohamedamin Abdullahi, Nassir Abdi Sheikh, Abeid Omar, Omar Abdihamid

## Abstract

**Background:** Cancer is an increasing public health challenge in Kenya, particularly in rural and underserved regions where surveillance systems and diagnostic capacity remain limited. Kilifi County, located along the Kenyan coast, lacks a population-based cancer registry, and data on the local cancer burden is not available. This study aimed to characterize the demographic distribution of patients, cancer burden in the county, and management of cancer cases diagnosed at Kilifi County Referral Hospital (KCRH) over ten years.

**Methods:** This retrospective study analyzed the patterns of cancer in Kilifi County using patient records from KCRH during the study period (January 1, 2014, to January 1, 2024).

**Results:** A total of 101 patients with cancer were identified, 58% female, with a mean age of 54 years. Most patients were from Kilifi North (47%), with a high proportion reporting no formal occupation (41%) or farming (26%). Esophageal and cervical cancers were the most common (18% each), followed by breast and prostate cancers (5% each), with other malignancies occurring infrequently. Histopathology was the primary diagnostic modality (88%). Staging data were incomplete in 70% of cases; among documented cases, the majority presented with advanced disease (21% stage IV). Due to limited local treatment capacity, approximately half of the patients were referred to tertiary centers for chemotherapy, radiotherapy, or surgery. At data cut-off, 43% had died, 25% were on treatment, and 29% were lost to follow-up, with only 2% completing treatment or under follow-up.

**Conclusions:** This study demonstrates a substantial cancer burden in Kilifi County and highlights critical gaps in diagnostic capacity, staging, and continuity of care. Strengthening cancer surveillance systems, expanding diagnostic and treatment infrastructure, and establishing a population-based cancer registry are essential to improving cancer outcomes and advancing equitable care in rural Kenya

## Background

Cancer represents a growing societal, public health, and economic challenge globally, accounting for nearly one in six deaths (16.8%) and over one in four deaths (22.8%) from non-communicable diseases (NCDs) (1). The global burden continues to rise, with the International Agency for Research on Cancer (IARC) approximating 20 million new cancer cases and 9.7 million cancer-related deaths in 2022 (2). Lung cancer remains the most diagnosed cancer worldwide, followed by breast and colorectal cancers, while breast cancer is the leading malignancy among women. Alarmingly, cancer-related mortality is projected to increase substantially, with low- and middle-income countries (LMICs) expected to bear the greatest share of this burden (3).

In Kenya, the cancer burden is rapidly increasing, reflecting broader epidemiological transitions driven by population growth, aging, and changing lifestyle risk factors (4). According to GLOBOCAN 2022 estimates, Kenya recorded approximately 44,700 new cancer cases and over 29,000 cancer-related deaths, with breast, cervical, prostate, and esophageal cancers among the most common (5). Despite this growing burden, cancer outcomes remain poor due to late-stage presentation, limited access to diagnostic and treatment services, and significant health system constraints. These challenges are compounded by structural, political, and financing barriers, including high out-of-pocket costs, limited oncology workforce capacity, and inequitable access to diagnostic capacity and care across regions (6). While national efforts such as the Kenya National Cancer Control Strategy (2023–2027) aim to strengthen prevention, early detection, and treatment services, gaps in workforce development, infrastructure, and public awareness persist (7). Therefore, strengthening cancer care delivery and building a sustainable oncology workforce remain urgent priorities in Kenya and similar LMIC settings.

Kilifi County, located in Kenya’s coastal region approximately 420 km southeast of Nairobi, has a population of 1.45 million as per the Kenya National Bureau of Statistics, 2019 (8). The county faces substantial socioeconomic challenges, including a poverty rate of 72% and low literacy levels, vast semi-arid areas which increase vulnerability to climate change and poor health outcomes (9). Despite the growing burden of cancer, no population-based cancer registry currently exists in the coastal region of Kenya, limiting the availability of reliable epidemiological data to inform local cancer control strategies. This cancer data gap constrains evidence-based planning for prevention, early detection, and resource allocation. Nationally, cancer surveillance capacity remains limited, with only two established population-based registries located in the capital Nairobi and Eldoret, although Machakos County has recently initiated a modern cancer registry (10). Therefore, strengthening subnational cancer registration systems is critical to improving data-driven cancer control efforts across Kenya.

This study aims to address this gap by retrospectively analyzing the clinicopathological characteristics of cancer cases diagnosed at Kilifi County Referral Hospital (KCRH) over ten years (January 1, 2014, to January 31, 2024). The findings will provide baseline data on cancer patterns and outcomes in Kilifi County, inform locally relevant prevention and treatment strategies, and contribute to strengthening subnational and national cancer surveillance systems.

## Methodology

### Study Design and Population

This retrospective study analyzed the patterns of cancer in Kilifi County using patient records from KCRH during the study period (January 1, 2014, to January 1, 2024).

### Data Sources

Data were obtained from the hospital’s Medical Records Department

### Inclusion and exclusion criteria

Patients with a documented cancer diagnosis in hospital records during the study period were included. Eligible records contained sufficient demographic and clinical information, including age, sex, residence, cancer type, stage at diagnosis, and treatment pathway. Patients with insufficient data were excluded.

### Data Analysis

Data were analyzed using SPSS version 27. Descriptive statistics were used to summarize patient characteristics, including cancer type, stage at diagnosis, age, sex, and residence. Categorical variables were reported as frequencies and percentages. Given the exploratory nature of the study, the lack of a cancer registry in the region, small sample size, the analysis was restricted to descriptive statistics.

### Ethical Approval

Ethical approval was obtained from the Meru University of Science and Technology Institutional Research Ethics Review Committee (MIRERC 001/2025). Permission to conduct the study was also granted by the Office of the Medical Superintendent at KCRH, which oversees compliance with institutional policies and guidelines.

## Results

A total of 101 patients with documented cancer diagnoses were identified over the study period. Of these, 58% (n=59/101) were female and 42% male (n=42/101) with a mean age at diagnosis of 54 years (SD). Geographically, most of the patients, 47% (n=47/101), resided in Kilifi North sub-county, followed by Ganze 19% (n=19/101) and Kilifi South 18% (n=18/101). Smaller proportions were from Kaloleni 5% (n=5/101), Malindi 4% (n=4/101), Rabai 4% (n=4/101), and Magarini 3% (n=3/101). Regarding socio-economic characteristics, 41% (n=41/101) of patients reported no formal occupation, while 26% (n=26/101) were engaged in farming. Smaller proportions reported involvement in business or trading, 9% (n=9/101). Additional demographic and socio-economic characteristics are presented in Table 1.

**Table 1.**
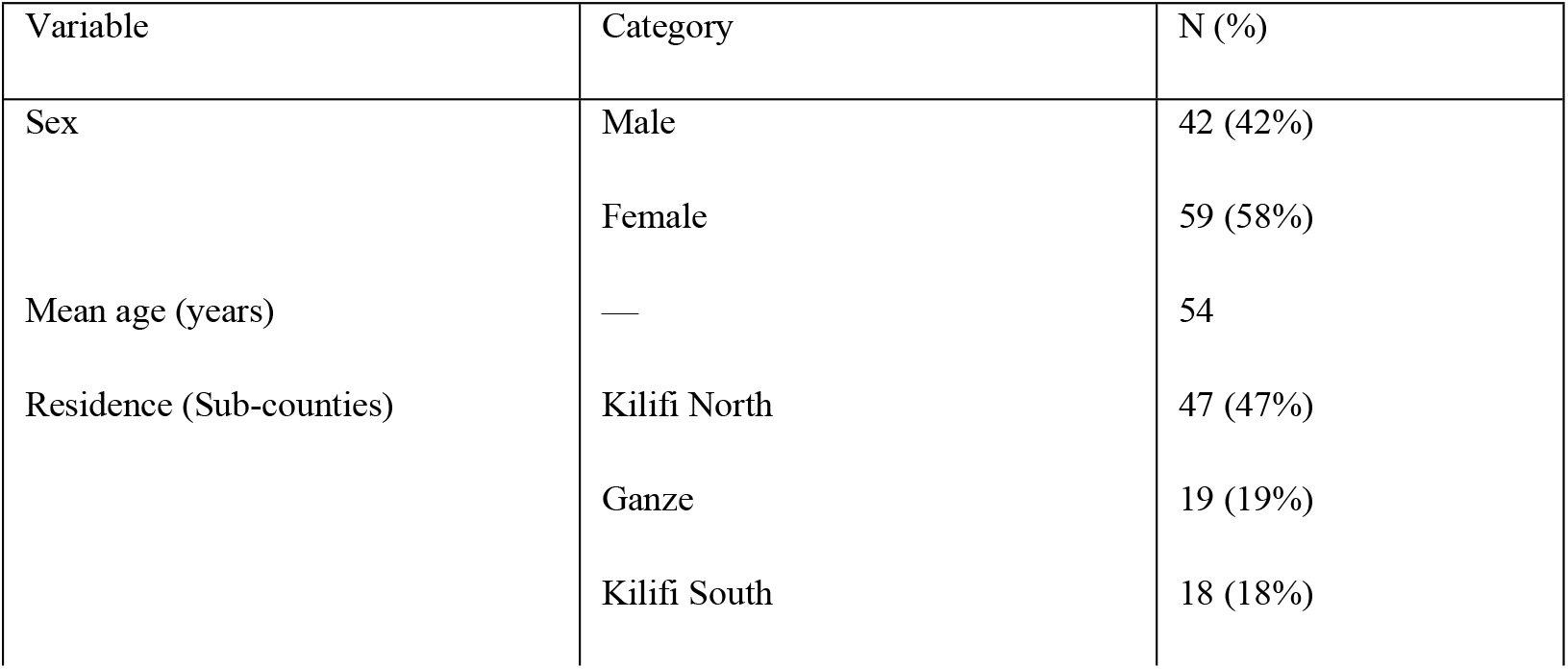
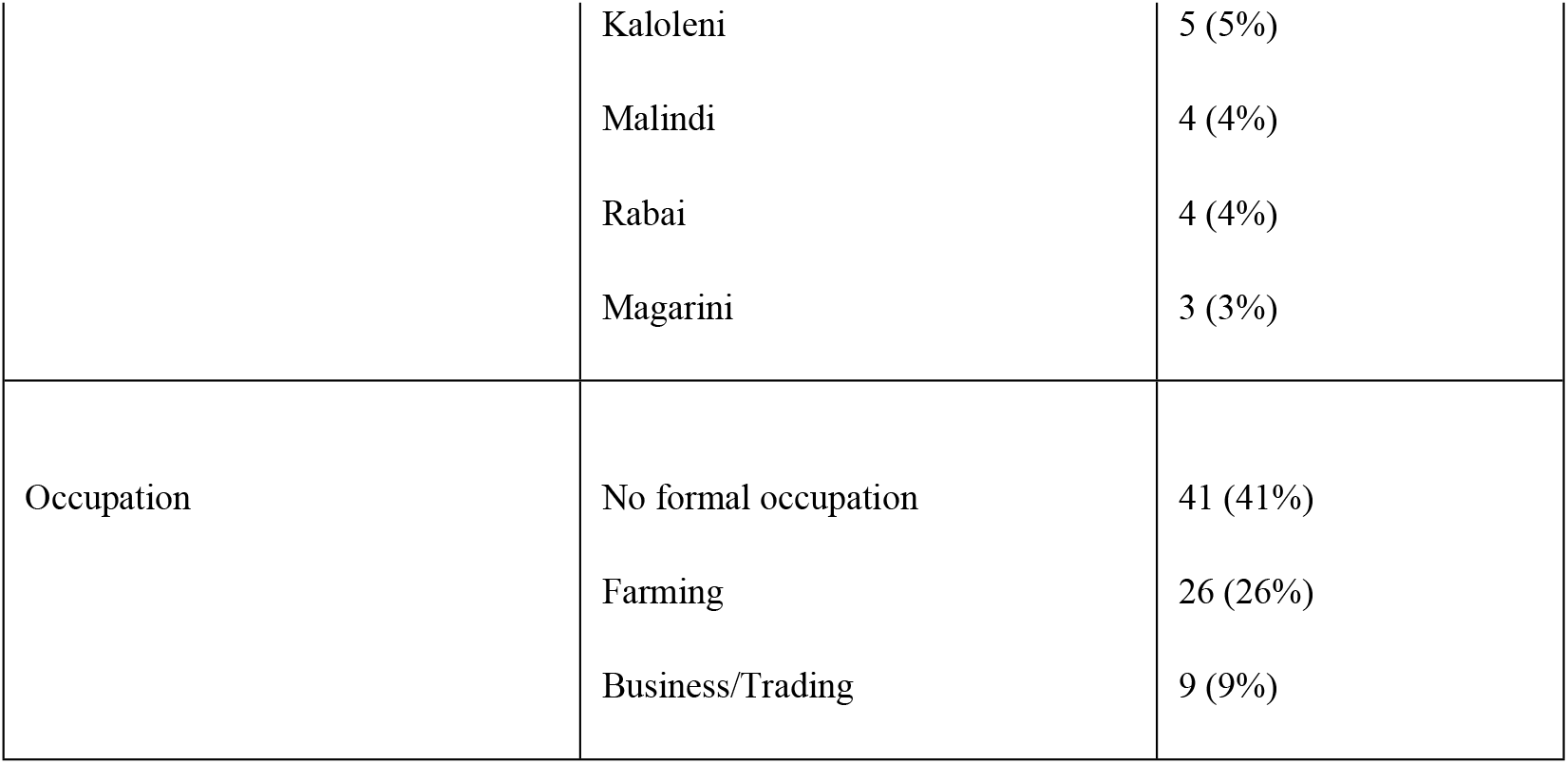
Baseline Characteristics of Study Participants (N = 101)

| Variable | Category | N (%) |
| --- | --- | --- |
| Sex | Male | 42 (42%) |
|  | Female | 59 (58%) |
| Mean age (years) | — | 54 |
| Residence (Sub-counties) | Kilifi North | 47 (47%) |
|  | Ganze | 19 (19%) |
|  | Kilifi South | 18 (18%) |
|  | Kaloleni | 5 (5%) |
|  | Malindi | 4 (4%) |
|  | Rabai | 4 (4%) |
|  | Magarini | 3 (3%) |
| Occupation | No formal occupation | 41 (41%) |
|  | Farming | 26 (26%) |
|  | Business/Trading | 9 (9%) |

Esophageal and cervical cancers were the most frequently documented cancers, each accounting for 18% (n=18/101) of cases. Breast and prostate cancers followed, each comprising 5% (n=5). Less common cancers included laryngeal cancer 3% (n=3/101), oropharyngeal 2% (n=2/101), and colorectal cancers 2% (n=2/101). A diverse range of other malignancies was identified in single cases (approximately 1% each, n=1/101), including hematological cancers, soft tissue sarcomas (osteosarcoma, Ewing’s sarcoma, myosarcoma, and endometrial sarcoma), Kaposi’s sarcoma, vulvar cancer, and dermatofibrosarcoma protuberans. The distribution of cancer types is shown in Figure 1.

**Figure 1.**
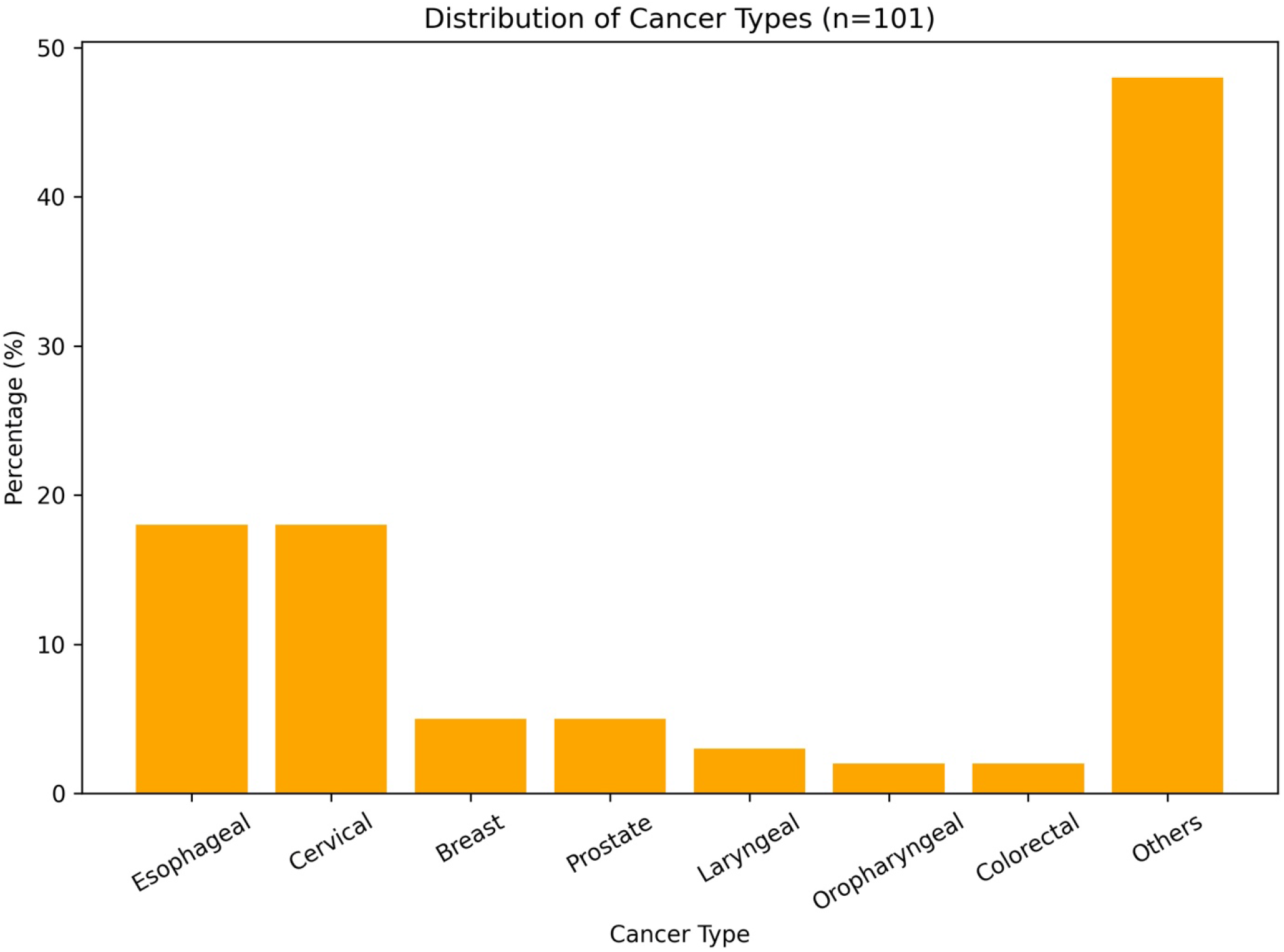
Distribution of cancer types among patients in Kilifi County (n=101)

Histopathology was the primary diagnostic modality, utilized in 88% (n=89/101) of cases, while 12% (n=12/101) were diagnosed radiologically. Cancer staging was incomplete in most cases, with 70% (n=71/101) lacking a documented stage at diagnosis. Among cases with available staging information, 2% (n=2/101) had stage II, 9% (n=9/101) had stage III disease, and 21% (n=22/101) had stage IV. Since KCRH did not have chemotherapy and radiotherapy services during the study period, approximately 50% (n=51/101) of patients were referred to regional tertiary care facilities, such as Coast General Teaching and Referral Hospital, Aga Khan Hospital in Mombasa, for treatment, including chemotherapy and radiotherapy, and would occasionally come back to KCRH for follow-up and for management of toxicity and supportive care. Among those referred, the majority received chemotherapy, alone or with concurrent radiotherapy, and surgery. At the data cut-off, 43% (n=43/101) had died, 25% (n=25/101) were on active treatment, and 29% (n=29/101) were lost to follow-up. Only 2% (n=2/101) had completed treatment or were in follow-up (Figure 2).

**Figure 2.**
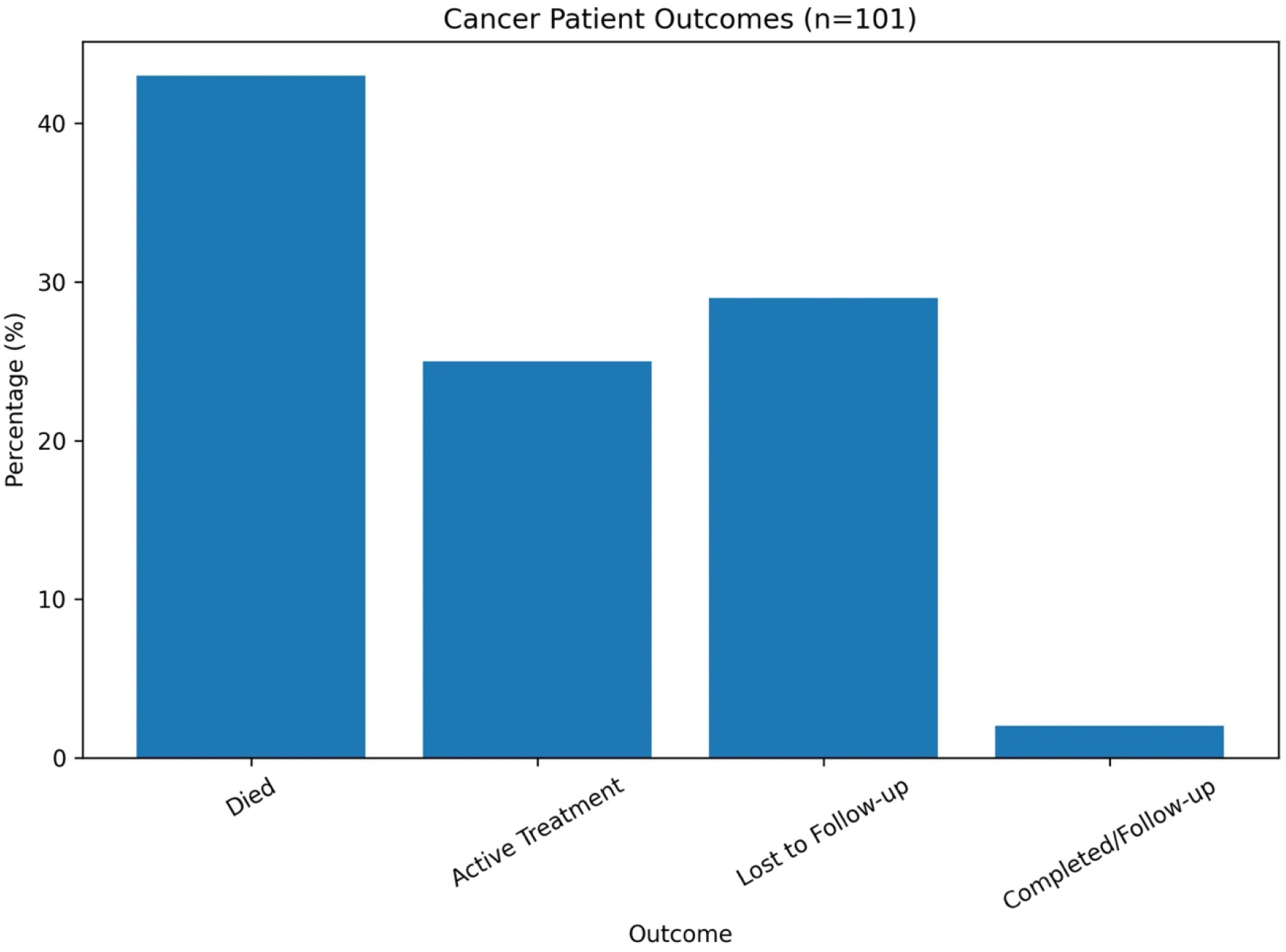
Distribution of patient outcomes among cancer cases in Kilifi County (n=101)

## Discussion

This study provides the first retrospective description of cancer burden at Kilifi County Referral Hospital (KCRH), a region with significant cancer burden but with unequal cancer infrastructure, despite recent efforts by the Kenyan government to set up a cancer center in the region. The study revealed an obvious socioeconomic vulnerability among cancer patients at KCRH, with nearly half of the patients lacking formal employment. This likely contributes to the lack of staging and treatment completion or follow-up as most patients could not afford these services, which were not readily available at the KCRH, forcing them to travel several kilometers away from their place of residence.

During the study period, esophageal and cervical cancers were the most documented cancers in Kilifi County, with significant diagnostic and follow-up limitations. These findings reflect broader regional cancer trends, especially cervical cancer in the coastal region but also highlight critical gaps in local cancer surveillance and care in Kenya (11, 12). Esophageal and cervical cancers accounted for 36% of all cases, consistent with patterns reported in other parts of Kenya and East Africa(13, 14). Cervical cancer is a leading cause of cancer-related incidence and mortality among women in Kenya, with an estimated 3,200 deaths in 2020 alone (15), coupled with inaccessible diagnostic modalities and poor uptake of cervical cancer screening and HPV vaccines(16). Uptake of HPV vaccination in Kenya has been sub-optimal, with only 33% of the targeted population receiving the first dose in 2020 and 16% returning for the 2nd dose (17). Esophageal cancer, on the other hand, is the third leading cancer-related mortality among both males and females in Kenya, according to the recent 2022 GLOBOCAN data which was published in 2024 (5).

The relatively low detection of breast and prostate cancers in our study, each comprising only 5%, may reflect underdiagnosis or access-related disparities rather than a true epidemiologic deviation from Kenya national cancer data (18). Breast is the number one common cancer among women in Kenya and the second leading cancer-related mortality, while prostate cancer is the most common cancer among Kenyan men and the fifth in cancer-related mortality in Kenya (5).

In the present study, histopathology was the primary diagnostic modality of cancer; however, most cases (70%) were unstaged, highlighting a significant gap in diagnostic evaluation and cancer staging capacity within the setting. This limited staging data underscores systemic weaknesses in diagnostic capacity and continuity of care, common in other resource-limited Kenyan counties (19). Nevertheless, the hospital has recently installed a CT scanner and a histopathology lab, which are expected to significantly improve the diagnostic capacity for cancer cases in the hospital and the county.

There was a high mortality rate (43%) and an extremely low treatment completion or follow-up rate. The significant proportion of patients referred elsewhere for care further emphasizes the constrained oncology infrastructure at the county level. Currently, the hospital is in the final stages of setting up a cancer center, which will significantly cut down the referrals to other cancer centers, thus improving treatment completion rates and follow-up.

None of the patients had a documented family history of cancer or awareness of hereditary cancer risk. This low awareness could be due to data omission and incomplete past medical history capture in the patient file, but it is consistent with prior findings from rural Kenya, where cancer literacy and screening uptake remain low, with one study reporting that most respondents (>50%) could only recognize one to two cancer types. In the same study, perceptions of survival from cancer were particularly pessimistic, with <70% recognizing early detection to improve survival outcomes (20).

The strength of our study is that it highlights the urgent and unmet need for a population-based cancer registry in Kilifi County, and investment in diagnostic and staging infrastructure, and targeted public health interventions to improve awareness, screening, and early detection. Strengthening local capacity and data systems will be essential to guide effective cancer control strategies and reduce the growing burden of non-communicable diseases in rural Kenya. The study also offers an important initial glimpse into the cancer burden in Kilifi County, a region that has long remained underrepresented in national cancer data. By analyzing the county’s cancer cases over a decade, we were able to identify meaningful patterns in cancer types, demographic characteristics, and challenges in cancer care pathways in a real-world setting. In doing so, this work begins to address a critical data gap and can inform local policy and resource planning, particularly in similar underserved rural areas.

That said, the study has several limitations. As a retrospective review based on hospital records, it reflects only those patients who accessed care at the facility and may not capture the full spectrum of cancer in the community. The small number of cases and the high proportion without staging information limit our ability to comment on disease progression and outcomes. Furthermore, many patients were referred to other institutions, and follow-up data were often incomplete, making it difficult to assess treatment completion or survival. Finally, the lack of information on behavioral or environmental risk factors, such as tobacco use, dietary habits, or infections, precludes a more nuanced understanding of cancer etiology in this setting.

## Conclusion

This study highlights a substantial cancer burden in Kilifi County, with esophageal and cervical cancers predominating, and underscores significant gaps in diagnosis, staging, and continuity of care. These findings reflect broader structural challenges in the delivery of cancer services in rural Kenyan settings. Despite the limitations of its retrospective, facility-based design, the study provides important baseline data from a previously underrepresented region, contributing to the evidence base for subnational cancer control planning. Strengthening diagnostic and staging capacity, improving clinical documentation, and establishing a population-based cancer registry in Kilifi will be critical to enhancing surveillance, informing resource allocation, and advancing equitable cancer care.

## Data Availability

All data available for free

